# Modelling the Impact of Obesity Reduction on the Prevalence of Hypertension in India: A Discrete-Event Microsimulation Approach

**DOI:** 10.1101/2024.01.24.24301738

**Authors:** Akif Mustafa

## Abstract

Obesity is one of the most significant risk factors of non-communicable diseases, disability, and premature death. Due to its profound impact on health, researchers have started classifying it as a disease rather than a mere abnormality. India, following the global trend, is experiencing a surge in obesity prevalence, posing a critical research question about the potential impact of obesity reduction on NCD incidence and related disorders. This study employs discrete-event dynamic microsimulation modelling to investigate how changes in BMI distribution in early years of life can influence the prevalence of hypertension, one of the most prevalent diseases in India. The microsimulation modelling approach enables the simulation of individual-level real-life behaviors and interactions within a given population. The model simulated the lives of 100,000 individuals aged 20 over the next 50 years till age 70. Baseline characteristics, prevalence rates, and transition probabilities were derived from diverse data sources, including Census 2011, the National Family Health Survey - V (NFHS-5), and the Longitudinal Aging Study in India (LASI, 2017-18). The study explores the impact of two scenarios on hypertension prevalence: (i) a one-unit reduction in mean BMI level at baseline, and (ii) a one-unit reduction in the standard deviation of BMI distribution at baseline. Results indicate that a one-unit reduction in mean BMI level at baseline could lead to a 5% reduction in hypertension prevalence at age 70, while a one-unit reduction in the standard deviation of BMI distribution at baseline could result in a 7.5% reduction. These findings underscore the importance of targeting children and adolescents with elevated BMI values to mitigate the later-life prevalence of hypertension. Additionally, the study highlights the significance of promoting the use of microsimulation modelling in health research in the Indian context.

## 1 Introduction

Obesity is linked to an elevated risk of nearly all noncommunicable diseases and disabilities. Conditions such as diabetes, hypertension, stroke, coronary heart diseases (CHDs), and even eye diseases like glaucoma have been found to be associated with obesity [1, 2, 3]. Not only NCDs, obesity has been found to significantly increase the risk of communicable diseases as well. Recently, several studies reported that obesity is correlated with a significantly higher risk of severe COVID-19 outcomes and mortality [4]. In light of these detrimental effects on health, researchers have begun to view obesity as a disease rather than a mere abnormality [5]. This perspective shift has been recognized by major health institutions such as the National Institutes of Health (USA) and the American Obesity Society, which now advocate for the treatment and prevention of obesity as a disease [6].

What is more concerning is that despite heightened awareness and information about the risks associated with obesity, its prevalence is on the rise globally, including in India. According to the World Health Organization (WHO), the worldwide incidence of obesity has nearly tripled since 1975. In 2016, over 1.9 billion adults (39%) were overweight, with over 650 million classified as obese (13%) [7]. These numbers are projected to increase even further in the future [7]. India is no exception to this public health issue. In fact, the incidence of over-weight and obesity is escalating at a pace surpassing the global average. Based on a recent study utilizing NFHS-5 (2019-21) data, the prevalence of abdominal obesity in the country stands at 40% among women and 12% among men [8]. However, when considering BMI criteria, 23% of women and 22.1% of men are categorized as overweight according to NFHS-5 data. Another study forecasts that the prevalence of overweight will more than double among Indian adults aged 20–69 years between 2010 and 2040, while the prevalence of obesity will triple [9]. These statistics not only underscore the immediate health concerns but also raise critical questions about the burden of morbidity in the coming decades.

From a policy perspective, it is crucial to understand the potential impact of reducing BMI levels on the overall burden of morbidity. Equally important is gaining insight into the most effective strategies and interventions that can yield optimal outcomes. One approach to achieve this is through the utilization of microsimulation models, which have been widely employed in various domains such as economic policy development (e.g., tax policies), urban planning, transportation planning, labor market studies, insurance and risk management, and energy and environmental policy.

Microsimulation is a technique that allows for the simulation of real-life behaviors and interactions within a given population. Unlike aggregate-level models that treat populations as homogeneous entities, microsimulation enables researchers to create virtual populations where each individual is a unique agent with distinct characteristics, behaviors, and attributes. In this type of simulation modeling, individuals can interact with one another, forming an artificial society that simulates a hypothetical population of interest. Researchers use this technique to examine how introducing a policy or intervention would impact the individuals in the population and shape the population dynamics. Using microsimulation modeling, this study examines how changes in obesity levels or alterations in BMI distribution can lead to a reduction in the prevalence of hypertension.

There are two primary reasons that contribute to the selection of hypertension as the outcome in this study. Firstly, hypertension is a serious concern within the Indian population and one of the most prevalent NCDs in the country. Secondly, the availability of micro-level data on blood pressure (both systolic and diastolic) at the national level is essential for defining the parameters in the microsimulation models used in this study.

Hypertension, also called as “silent killer,” earns its name because many individuals with hypertension remain unaware of the issue. For a substantial duration, it may exhibit no signs or symptoms, silently compromising individuals’ health. According to the World Health Organization (WHO), approximately 1.28 billion adults aged 30–79 years worldwide suffer from hypertension [10]. A study reported that 8.5 million deaths in 2015 were associated with high blood pressure [11]. In India, hypertension is widespread, with NFHS-5 indicating that 24% of men and 21% of women aged 15 and above have hypertension. Considering the imminent rapid population aging in India, with a substantial number entering older age groups, this prevalence is anticipated to rise further.

Given that obesity is a significant risk factor for hypertension, and research suggests that obesity alone can induce hypertension [12, 13], this study, using dynamic discrete-event microsimulation modeling, aims to explore how changes in BMI distribution at early stages of life can result in reduction of hypertension incidence and prevalence.

In addition to investigating the impact of changes in BMI distribution on hypertension prevalence, this article holds significant value in promoting use of microsimulation modelling in health research in India. While this methodology is widely utilized in developed countries, its potential to analyze the impacts of existing and proposed social and economic policies at both micro and macro levels remains largely unexploited in research and policy formations in India. The article aims to raise awareness about the importance of microsimulation modeling in the Indian context, not only in the domain of obesity but across all areas of health research. By highlighting the potential benefits and applications of this modeling approach, the author aims to encourage its wider adoption and utilization among researchers and policymakers.

## 2 Methods

### Model Structure

In this study, we utilized a discrete event dynamic microsimulation model to analyze the progression of 100,000 individuals (agents) from age 20 to 70 over a span of 50 years. Due to data limitations and the complexity of the relationship between BMI and blood pressure at inital and later stages of life, the model does not simulate individuals from birth to death. The model initially explores the simulated outcome (hypertension) over the course of 50 years if the individuals at age 20 experience the current patterns of BMI, blood pressure, and the relationship between the two. The model further investigates how slight reduction in mean and standard deviation of BMI distribution in early years (age 20 in this study) can impact hypertension rates. Further characteristics of the model are described below.

### Baseline Population

The study began with a baseline population of 100,000 individuals, who were randomly assigned to one of four categories: urban male, urban female, rural male, and rural female. The probability of an individual being in a rural area was calculated to be 0.692, while the probability of being in an urban area was 0.308. Within the rural area, the probability of being male was 0.517, and in the urban area, it was 0.515. These probabilities were obtained from the 2011 census data, ensuring the accurate representation of the population distribution in the simulation.

### Mortality Pattern

We obtained the probabilities of death from the abridged life table of India 2016-20, which was published by the Sample Registration System (SRS) of the Office of Registrar General & Census Commissioner. The SRS releases annual abridged life tables that provide information on life expectancy for different age groups, categorized by sex and residence (urban or rural). In our study, we required the probability of death for each year from age 20 to 69. However, the available data only provided probabilities for five-year age groups, such as 0-5, 5-10, and so on. To address this, we used an interpolation method in Excel. In excel, we combined OFFSET, MATCH, and FORECAST functions to perform the interpolation. The method extracts data points from the 5-year age group values that straddle the target year and employs interpolation to estimate the probability of death for that specific year. The method we used was different from the simple linear interpolation method which can be employed using the Forecast function. Instead, our method assigns greater weight to the years closest to the target year when calculating the interpolated probability of death. This interpolation approach enhances the accuracy of our estimates by placing more significance on adjacent years in the calculation process. Year-wise probabilities of death in the four groups are presneted in Figure 1.

**Figure 1:**
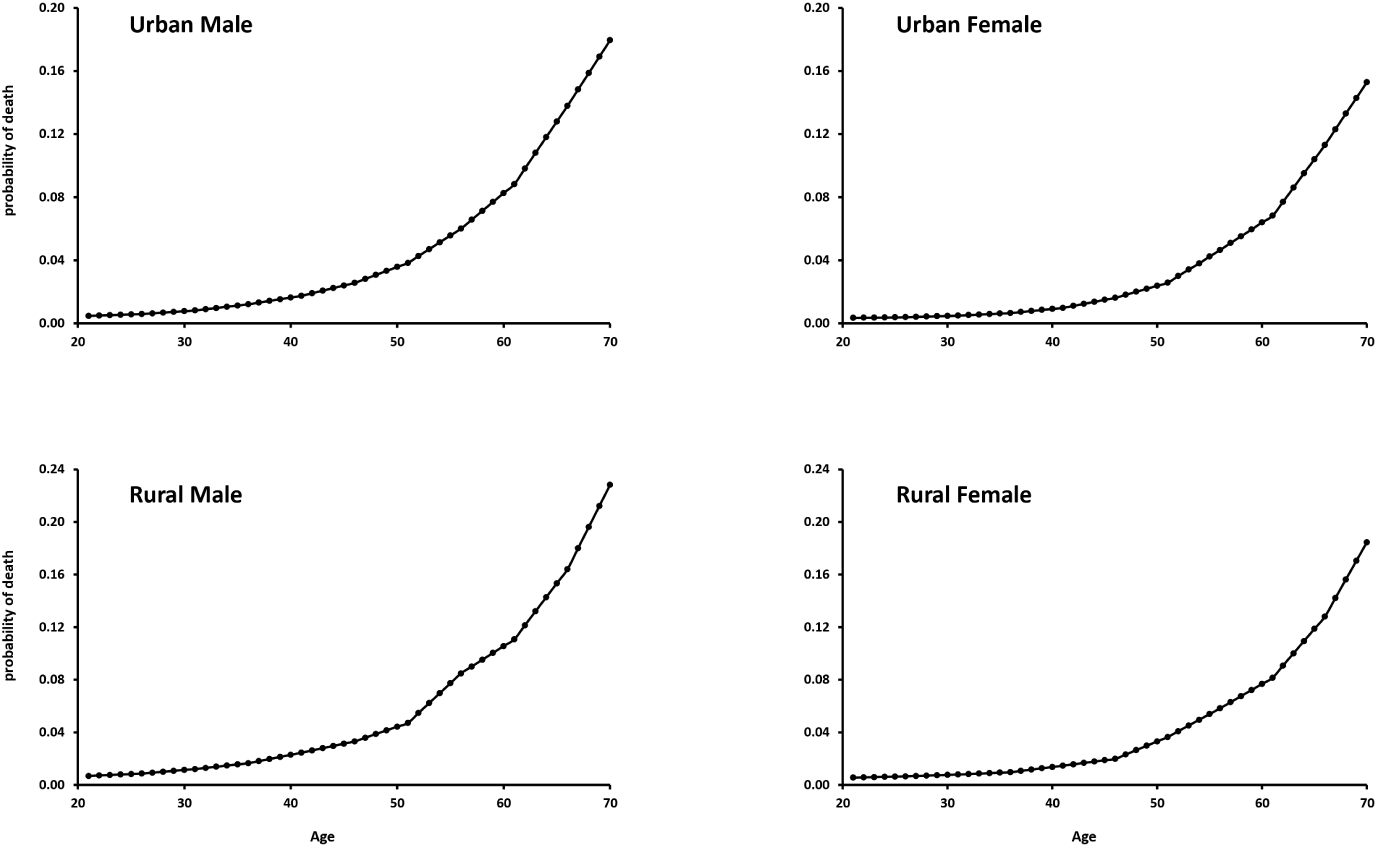
Distribution of probability of death by age.

### BMI and Blood Pressure data

In order to establish parameters for the baseline BMI data and estimate how BMI changes with age, we relied on data from the fifth round of the National Family Health Survey (NFHS-5) and the Longitudinal Ageing Study in India (LASI, 2017-18). We chose to use these two datasets because they not only provide information on BMI but also include data on systolic and diastolic blood pressure at the individual level. This allows us to establish the relationship between BMI and blood pressure that will be used in the microsimulation.

However, due to the specific age range requirements (20 to 70 years), it was necessary to employ two datasets rather than one. The NFHS dataset provides data for women aged 15 to 49 and men aged 15 to 54, while the LASI dataset covers the required age range from 45 to 80+ years. Although the LASI dataset does contain some data for individuals below the age of 45, the sample size for those ages was not sufficient. Therefore, the utilization of both datasets was imperative to ensure comprehensive coverage of the desired age range.

We gathered the necessary data from both sources and appended them into one dataset. Specifically, we obtained data for individuals under the age of 45 from the NFHS dataset, data for individuals over 50 years old from the LASI dataset, and data for individuals aged 45 to 49 from both datasets. It’s important to note that since the two datasets were collected using different sampling procedures and at different time points, there may be variations in the estimates between the samples. To assess this, we compared the average BMI levels specifically for the age group of 45 to 49 (as both datasets provide data for this age range). We observed only a slight difference in the BMI estimates.

### BMI Input Parameters

To define the baseline BMI and to estimate how BMI changes overtime we had to smoothen the data. We calculated mean BMI by age in all the four sex-residence groups. Then we plotted a local polynomial smooth between BMI and age and stored the smoothened BMI levels along with 95% confidence interval values. The plots of local polynomial smooth are shown in Figure 2.

**Figure 2:**
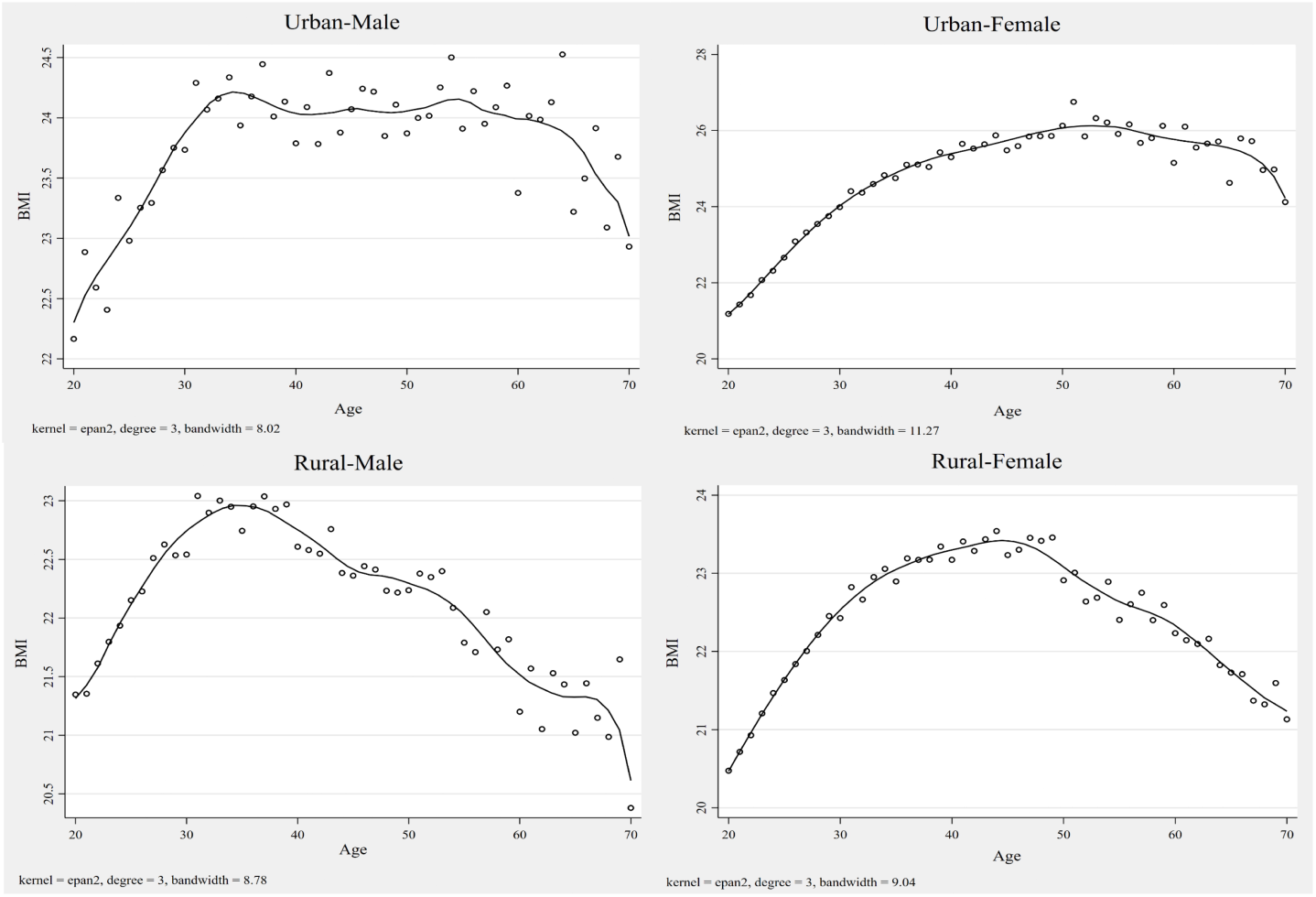
Local Polynomial Smooth of age and BMI.

Figure 3 illustrates the kernel density plot of BMI in the observed data. It is evident from the plot that the distribution of BMI is skewed to the right and not symmetrical. It appears to follow more of a log-normal distribution rather than normal distribution. Therefore, we distributed the BMI values in the baseline age (age 20) of the simulation population using a lognormal distribution. The mean value for BMI was derived from the smoothed mean value at age 20, while the standard deviation was obtained from the observed data. The mean BMI value, along with the standard deviation for the baseline BMI simulation, is provided below in Table 1.

**Figure 3:**
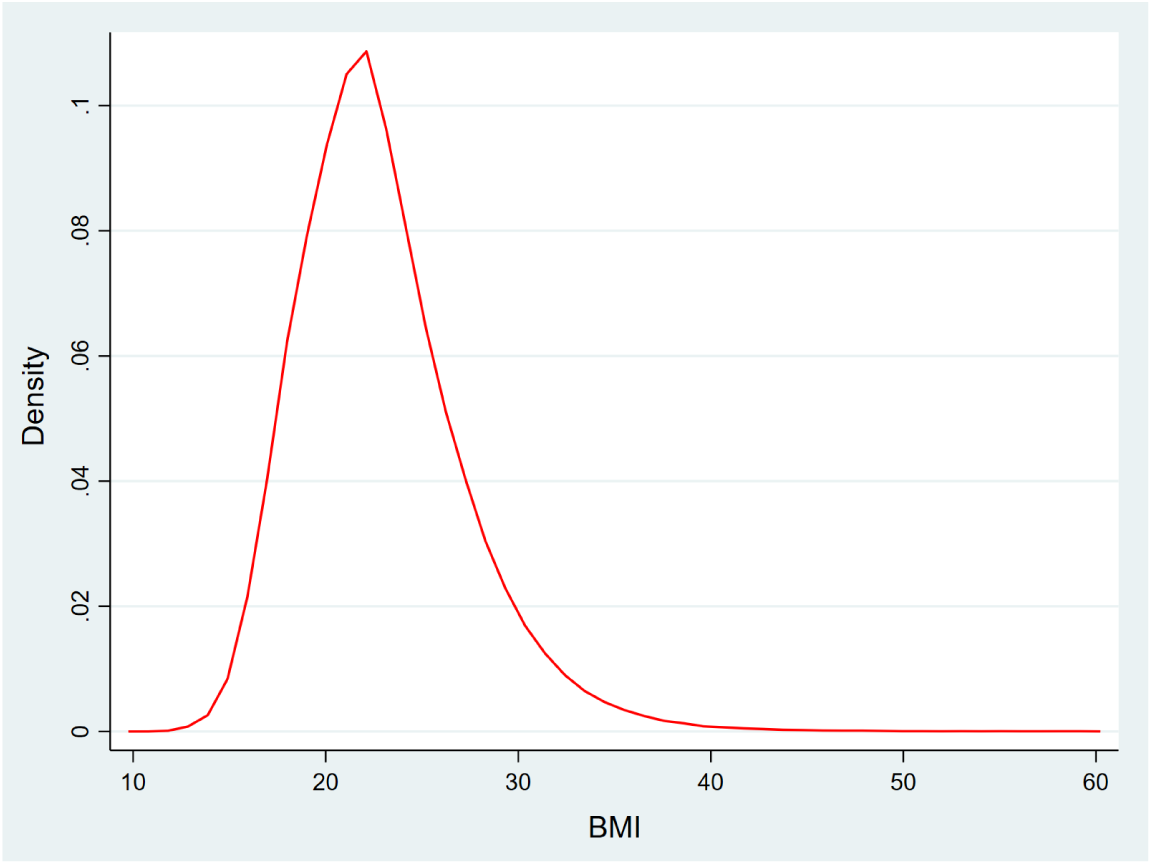
Kernel Density Plot of BMI in the observed data.

**Table 1:** BMI Input parameter for baseline distribution.

|  | Mean BMI | SD |
| --- | --- | --- |
| Urban Male | 22.30 | 4.23 |
| Urban Female | 21.17 | 5.12 |
| Rural Male | 21.31 | 4.43 |
| Rural Female | 21.05 | 4.35 |

Defining the annual evolution of BMI in the microsimulation was a crucial and complex task. We opted for a lagged value approach, which means that an individual’s BMI at a particular age would be influenced by their BMI from the previous year. This approach introduced a memory element to our model, distinguishing it from a simple Markov model where values are independent of lagged values, and lack memory. By considering the influence of past BMI values on present ones, our model captures the dynamic nature of BMI changes over time. This approach ensures that our findings align closely with real-world observations and provides a more accurate representation of how BMI evolves in individuals.

To better understand how we defined the BMI evolution in our model, let’s consider an example. Suppose we have an urban-male individual with a BMI value of 24.2 at age 20, which was randomly assigned using log-normal distribution. To determine how much the BMI will increase or decrease in the next year, we refer to the local polynomial smooth for urban males. At age 20, the smoothed BMI value for urban-males is 22.30. For the next year (age 21), the smoothed BMI value is 22.53, with a 95% confidence interval ranging from 22.18 to 22.87. By calculating the differences between the BMI value at age 20 and the lower and upper bounds of the 95% confidence interval at age 21, we obtain:

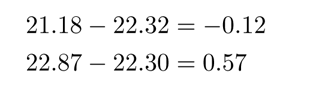

Therefore, for our individual with a BMI of 24.2 at age 20, the BMI at age 21 will fall within the range of (24.2 - 0.12 = 24.08) to (24.2 + 0.57 = 24.77). In our model, we introduce more realism and natural variation by using a uniform distribution to randomly assign a BMI value between the two values. In our example, the uniform distribution will assign any value randomly between 24.08 to 24.77 as a BMI value at age 21. In this case, the individual’s randomly assigned BMI at age 21 is 24.63.

To determine the BMI at age 22, we repeat the process by referring to the smoothed data. At age 21, the smoothed BMI value is 22.53, and the 95% confidence interval for age 22 ranges from 22.36 to 23.01. The differences between these values are:

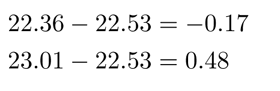

Thus, for our individual, the BMI value at age 22 will be within the range of (24.63 - 0.17 = 24.46) to (24.63 + 0.48 = 25.11). Once again, we use a uniform distribution to assign a randomly generated BMI value between 24.46 and 25.11 for age 22. This process is repeated for all individuals till age 70. The difference values generate using the local polynomial used which are used in defining the annual evolution of the BMI are presented in appendix Table A1.

### Blood Pressure parameters

In our model, we determined the systolic and diastolic blood pressure based on the individual’s BMI and age. For each sex-residence group, we estimated the relationship between blood pressure (both systolic and diastolic) and BMI and age using linear regression. To account for the non-linear nature of the relationship between BMI and blood pressure, we included the squared term of BMI in the regression equation. The regression equation had the following form:

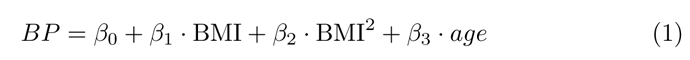

The equations of association for blood pressure and BMI can be accessed from appendix Table A2.

In our simulation model, an individual’s blood pressure was randomly assigned within the range of the lower and upper estimates of the equation using a uniform distribution. To illustrate this, let’s consider an example of a 35-year-old Rural-Female with a BMI of 23.6. According to the equation, the point estimate of her systolic blood pressure would be:

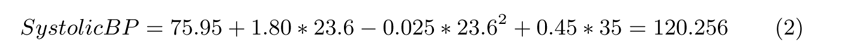

However, to add more dynamism and realism to our model, we decided to randomly assign the blood pressure value within the 95% confidence interval of the equation. The lower bound calculation yields:

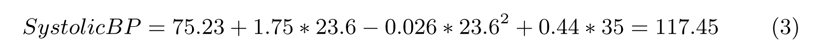

And the upper bound calculation yields:

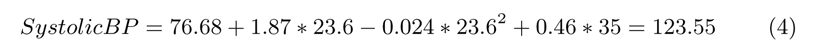

Therefore, the individual’s systolic blood pressure will be a randomly generated number between 117.45 and 123.55, assigned using a uniform distribution. By adopting this approach, our model achieves greater realism and dynamism, reflecting the inherent natural individual to individual variability. Let me explain how this approach is better. Suppose we adopted a deterministic approach, where all rural females aged 35 with a BMI of 23.6 would have a systolic blood pressure of 120.256 based on the point estimate of the equation. However, such an approach would oversimplify the complexity of real-life scenarios and overlook the variations that naturally exist among individuals. In contrast, our chosen approach introduces natural variability into the simulation model while still adhering to the equation of association. For instance, if there are multiple rural women aged 35 with a BMI of 23.6, they would each have a different systolic blood pressure within the 95% range of the equation. By incorporating this natural variability into our model, we create a more realistic simulation that better reflects the diversity and uniqueness of individuals.

## 3 Results

This section presents the outcomes of the initial baseline microsimulation model, which is built upon the original parameters derived from the observed data. While the microsimulation model simulates a cohort of individuals from age 20 to 70, the results are interpreted under the assumption that the cohort experiences of individuals represent a cross-sectional snapshot of a population at a given point in time. At age 20, the distribution of the individuals in the four sex-residence is presented in Table 2.

**Table 2:** Distribution of individuals at baseline age.

|  | n | % |
| --- | --- | --- |
| Urban-Male | 16,927 | 16.93 |
| Urban-Female | 17,337 | 17.34 |
| Rural-Male | 32,863 | 32.86 |
| Rural-Female | 32,873 | 32.87 |
| Total Population | 100,000 | 100.00 |

The distribution of the alive/death status of individuals at age 70 is shown in Table 3.

**Table 3:** Distribution of Individuals alive and dead at age 70.

|  | Alive<br>(%) | Dead<br>(%) |
| --- | --- | --- |
| Urban-Male | 1439 (8.50) | 15488 (91.50) |
| Urban-Female | 2560 (14.77) | 14777 (85.23) |
| Rural-Male | 1297 (3.95) | 31566 (96.05) |
| Rural-Female | 3115 (9.48) | 29758 (90.52) |
| Total Population | 8,411 (8.41) | 91589 (91.59) |

From the table, we can see that out of the total 100,000 individuals who were alive at age 20, approximately 91.6% died by the time they reached age 70. Among the different sex-residence groups, 8.5% of urban males, 14.8% of urban females, 3.9% of rural males, and 9.5% of rural females were able to reach age 70.

Figure 4 shows the mortality pattern in the four sex-residence group in the simulation model. The graph plots proportion alive on y-axis and age on x-axis.

**Figure 4:**
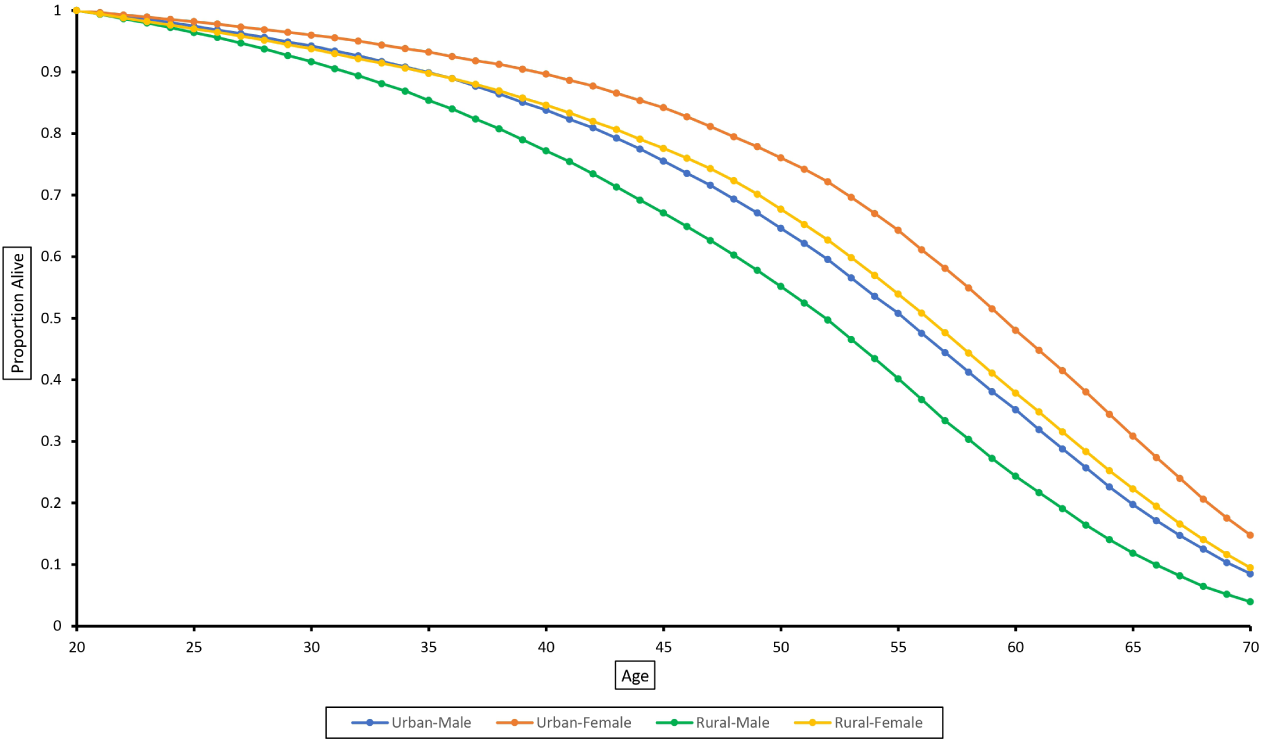
Mortality pattern in the simulated population by sex-residence group.

Figure 5 illustrates the density plot of the simulated BMI for the four distinct groups. It can be observed from the plot that the BMI distribution for urban individuals exhibits a broader spread towards the higher values on the right side of the plot, in comparison to the distribution for rural individuals. Additionally, the peak of the BMI distribution for urban individuals is situated at higher values, closely resembling the observed BMI distribution in the NFHS and LASI data.

**Figure 5:**
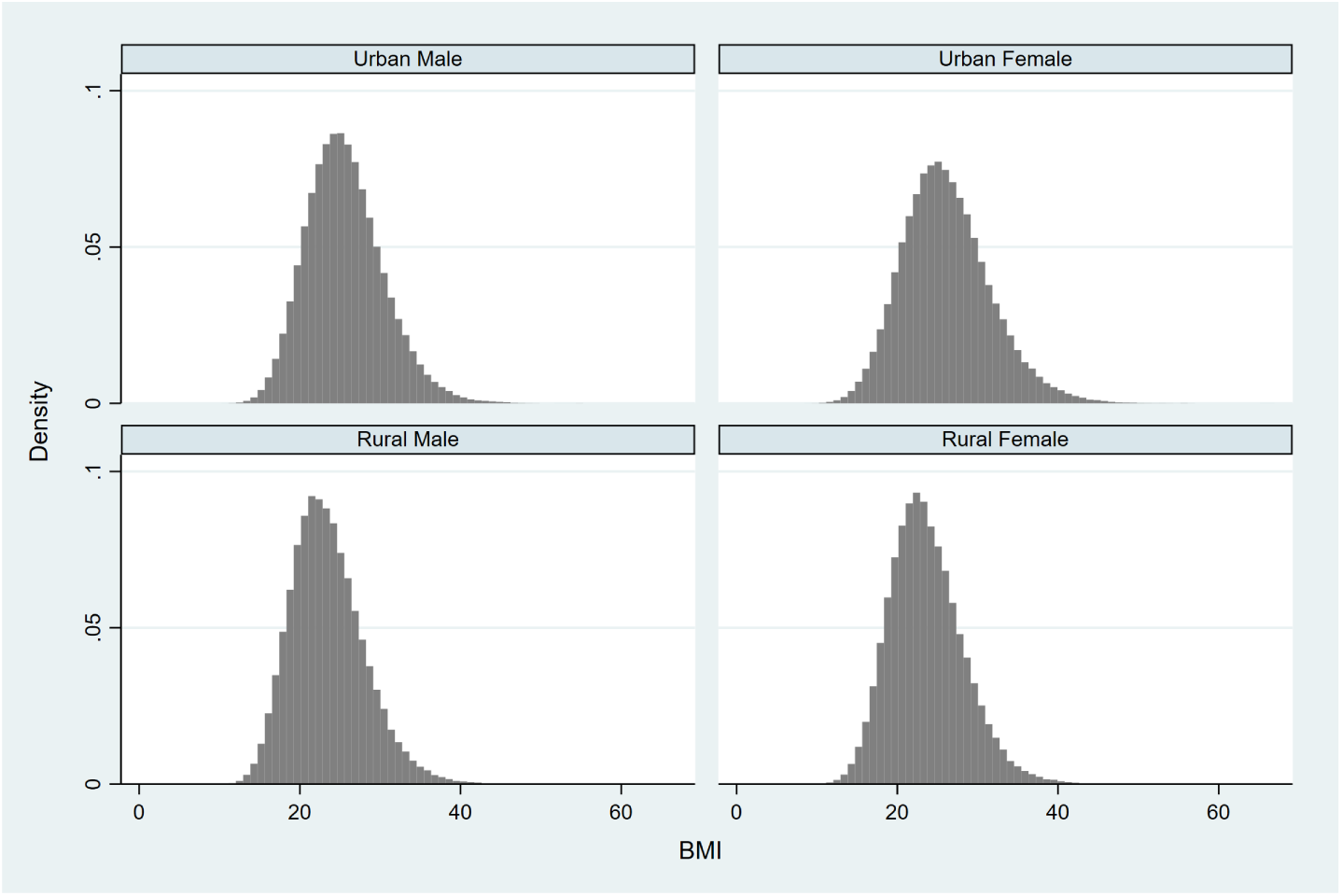
Distribution of BMI in the simulated population by sex-residence groups.

Figure 6 illustrates the progression of obesity and hypertension prevalence with age in our simulation model. Upon observing the figure, it becomes evident that the prevalence of both hypertension and obesity rises as individuals age. However, there are distinct patterns worth noting. The prevalence of obesity displays a modest decline after the age of 60. This can be attributed to factors such as mortality and the natural decrease in BMI that occurs with age. In contrast, the prevalence of hypertension does not exhibit any decline throughout the age range. As a chronic condition, hypertension persists and continues to affect individuals, resulting in a sustained increase in prevalence. Notably, after the age of 55, the prevalence of hypertension experiences a significant rise.

**Figure 6:**
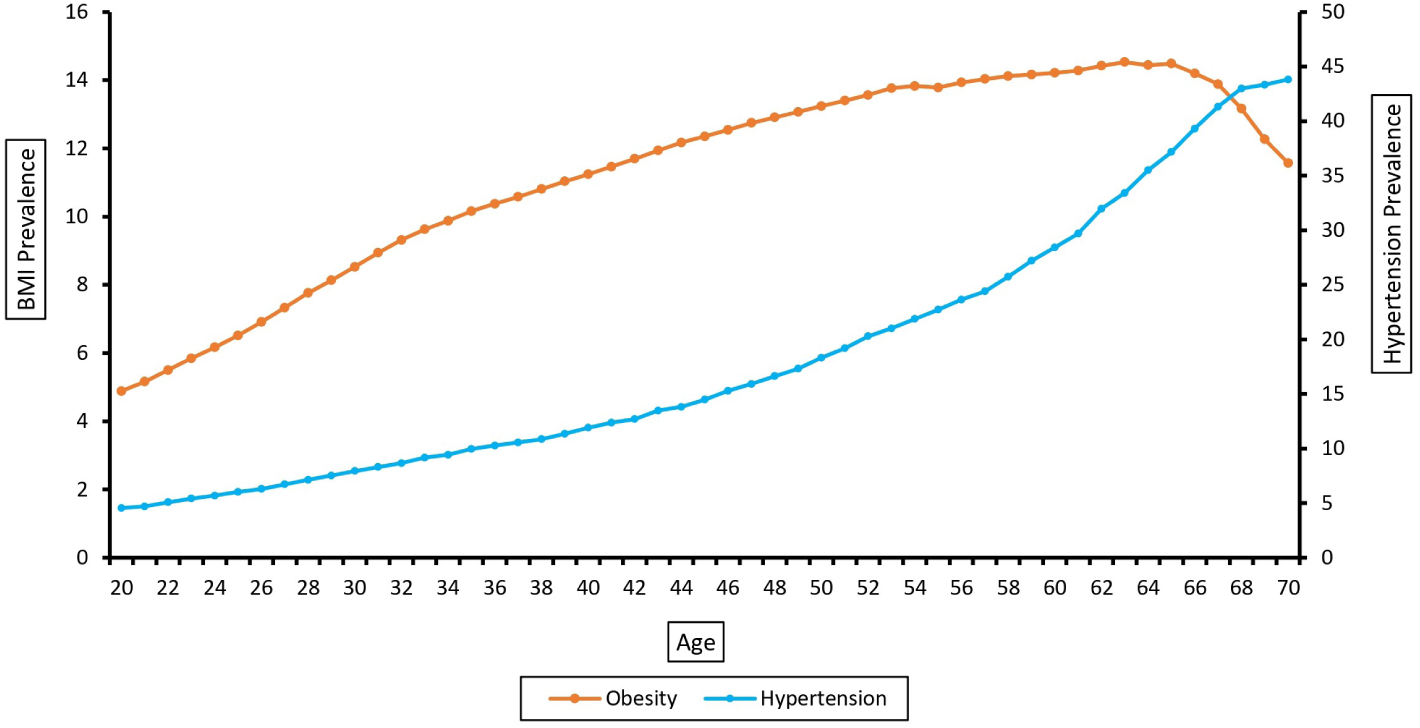
Progression of obesity and hypertension with age in the baseline simulation.

**Figure 7:**
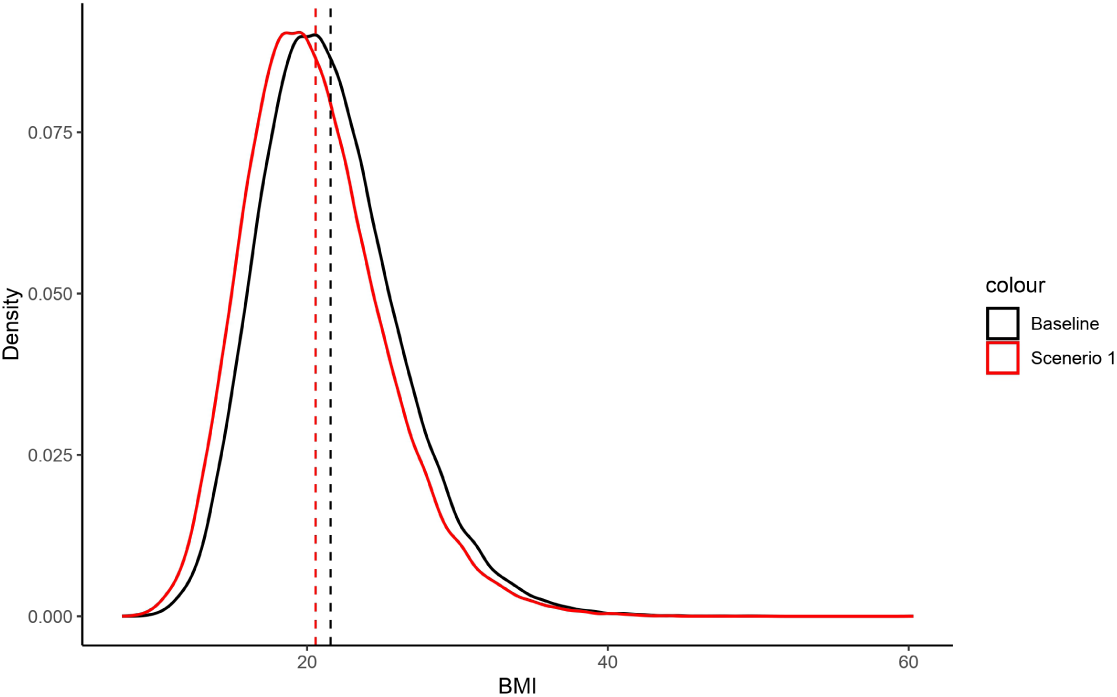
BMI distribution at age 20 under baseline and Scenario 1 simulation.

Table 4: presents the prevalence of obesity and hypertension in the simulation model, categorized by sex-residence groups and age. As expected, the highest prevalence of obesity was observed among urban females, reaching 17.41%, while the lowest prevalence was found among rural males, at 7.72%. The overall prevalence of obesity in the simulated population was determined to be 8.27%. At age 70 the prevalence of obesity was found to be 11.35%. This suggests that if the current population aged 20 years continues to follow the current BMI patterns, the projected prevalence of obesity among them at age 70 would likely be 11.35%.

**Table 4:** Prevalence of Obesity and Hypertension in the simulated population by age group.

|  | Urban<br>Male | Urban<br>Female | Rural<br>Male | Rural<br>Female | Total<br>Population |
| --- | --- | --- | --- | --- | --- |
| **Obesity** |  |  |  |  |  |
| Age Group |  |  |  |  |  |
| 20-29 | 7.46 | 9.01 | 5.86 | 5.21 | 6.40 |
| 30-39 | 11.87 | 15.85 | 7.74 | 7.68 | 9.90 |
| 40-49 | 15.15 | 19.31 | 8.81 | 10.11 | 12.17 |
| 50-59 | 17.60 | 20.90 | 9.77 | 9.67 | 13.73 |
| 60-70 | 18.78 | 20.10 | 8.31 | 8.87 | 13.87 |
| at age 70 | 18.67 | 16.41 | 6.77 | 7.25 | 11.35 |
| All | 13.19 | 17.41 | 7.72 | 8.11 | 10.27 |
| **Hypertension** |  |  |  |  |  |
| Age Group |  |  |  |  |  |
| 20-29 | 13.40 | 7.11 | 4.13 | 2.32 | 5.32 |
| 30-39 | 23.70 | 9.19 | 8.55 | 4.41 | 9.61 |
| 40-49 | 34.17 | 15.11 | 12.85 | 8.19 | 14.27 |
| 50-59 | 46.55 | 20.19 | 16.71 | 12.11 | 21.98 |
| 60-70 | 51.93 | 39.04 | 30.18 | 19.21 | 37.85 |
| at age 70 | 57.73 | 49.71 | 33.55 | 28.11 | 43.81 |
| All | 31.11 | 22.13 | 15.71 | 10.19 | 19.17 |

In contrast to obesity, the prevalence of hypertension displayed a different pattern. Among the sex-residence groups, urban males exhibited the highest prevalence of hypertension, reaching 31.11%, while rural females had the lowest prevalence at 10.19%. It is noteworthy that the prevalence of hypertension was relatively low in the younger age groups (20-29 and 30-39), but experienced a sharp increase in the later age groups. For instance, among urban males aged 50-59, the prevalence of hypertension was recorded at 46.55%. Furthermore, at age 70, the prevalence of hypertension in the simulated population was found to be 43.81%. This indicates that if a cohort of individuals aged 20 years were to follow the current pattern and relationship between BMI, age, and hypertension, the projected prevalence of hypertension among them at age 70 would be approximately 43.8%.

### Scenario Testing

In our scenario testing, we investigated the effects of modifying the baseline BMI distribution in two distinct scenarios.

**Scenario 1**: In the first scenario, we lowered the mean BMI for the baseline year (age 20) by one unit while maintaining the same standard deviation as in the first simulation across all four groups. Parameters for the Scenario-1 are presented in Table 5.

**Table 5:** BMI parameters for baseline year (age 20) in First simulation and Scenario-1 simulation.

|  | First Simulation<br>(Baseline) | Scenario 1 | SD<br>(same in both) |
| --- | --- | --- | --- |
| Urban-Male | 22.30 | 21.30 | 4.23 |
| Urban-Female | 21.17 | 20.17 | 5.12 |
| Rural-Male | 21.31 | 20.31 | 4.43 |
| Rural-Female | 21.05 | 20.05 | 4.35 |

The objective of this exercise was to assess the impact on hypertension prevalence when the average BMI of individuals at age 20 decreases by one unit. Figure-7 illustrates the change in the distribution of BMI at age 20 under the two situations (baseline and scenario 1 simulation).

**Scenario 2**: In the second scenario, we decreased the standard deviation for baseline BMI (age 20) by one unit in all four groups while keeping the mean BMI the same as in the first simulation (baseline). This scenario is comparatively more significant than the first because here we are reducing individuals at extreme BMI values without altering the mean. In the first scenario, we shifted the mean to the left by one unit, causing some individuals to transition from obese to non-obese status. However, it also led to some individuals becoming underweight due to the entire distribution shifting leftward. On the other hand, in the second scenario, by reducing the standard deviation, we are decreasing individuals from both extreme values, obese and underweight. Parameters for Scenario-2 are presented in Table 6.

**Table 6:**
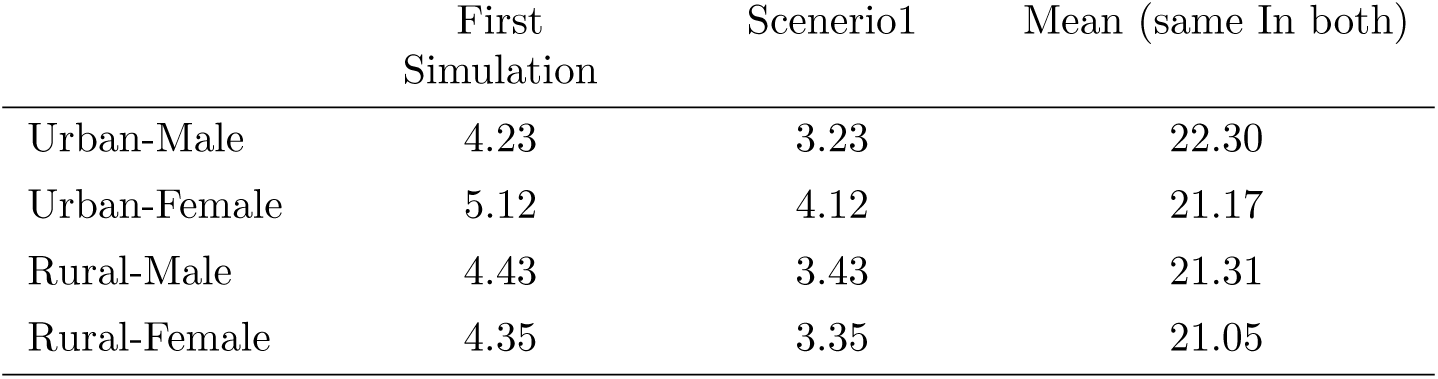
Standard deviation of BMI distribution for baseline year (age 20) in First simulation and Scenario-2 simulation.

The two scenarios suggest two distinct policy approaches. Scenario 1, which focuses on reducing the average BMI at the baseline age (age 20), suggests an intervention that is applicable to the entire population, such as implementing food package warnings and increasing the prices of sugary products. On the other hand, Scenario 2, which aims to reduce the standard deviation of BMI distribution, points towards a targeted approach where only individuals with higher BMI levels are selected for intervention. For instance, implementing a physical exercise and diet intervention specifically for obese adolescents.

Figure 8 shows the change in BMI distribution at baseline age due to oneunit decrease in the standard deviation of BMI.

**Figure 8:**
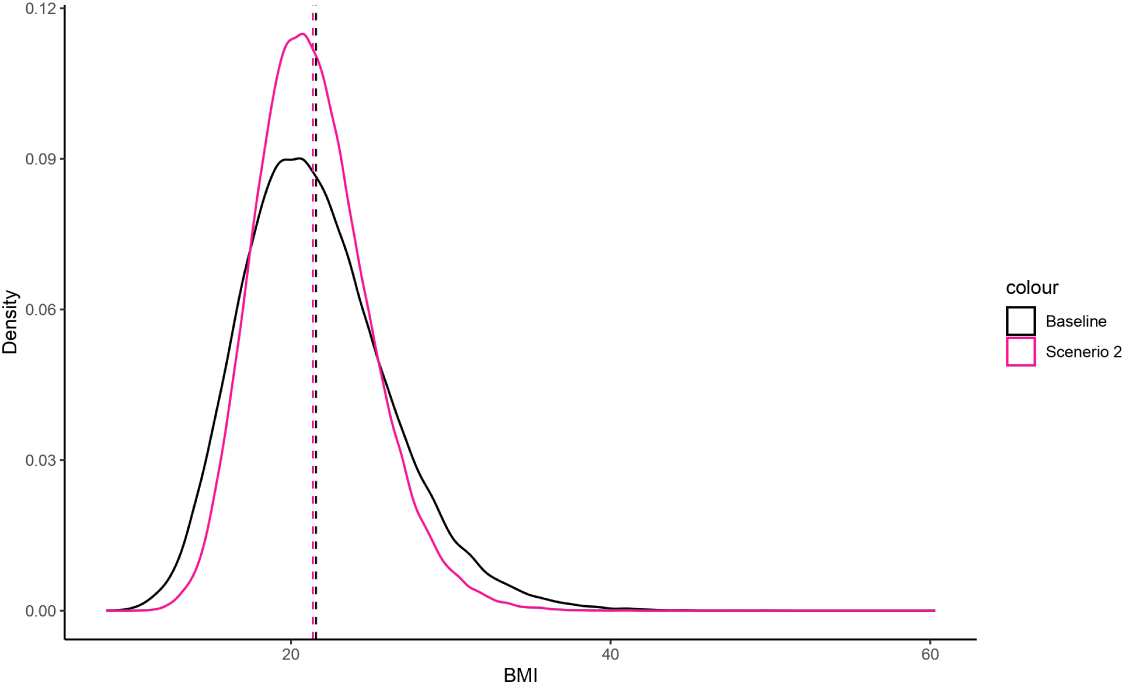
BMI distribution at age 20 under baseline and Scenario 2 simulations.

Table-7 provides an overview of BMI prevalence within the simulated population across three scenarios: baseline (first simulation), Scenario 1, and Scenario 2. As previously mentioned, in the baseline simulation, the obesity prevalence was 10.27%. However, in Scenario 1, this prevalence decreased to 7.79%. Furthermore, in Scenario 2, the obesity prevalence decreased even further to 6.88%.

**Table 7:** Prevalence of obesity under baseline, scenario 1 and scenario 2 simulations.

|  | Baseline | Scenerio1 | Scenerio2 |
| --- | --- | --- | --- |
| Urban-Male | 13.19 | 11.32 | 10.61 |
| Urban-Female | 17.41 | 14.23 | 13.11 |
| Rural-Male | 7.72 | 5.83 | 4.41 |
| Rural-Female | 8.11 | 6.31 | 5.76 |
| Total Population | 10.27 | 7.79 | 6.88 |

Figure-9 illustrates the density plot of systolic and diastolic blood pressure in the simulated population under the three scenarios. The graph shows that under Scenario 1, the distribution of both systolic and diastolic blood pressure shifted slightly to the left compared to baseline simulation. Under Scenario 2, not only did the distribution shift to the left, but it also became narrower, indicating a reduced spread.

**Figure 9:**
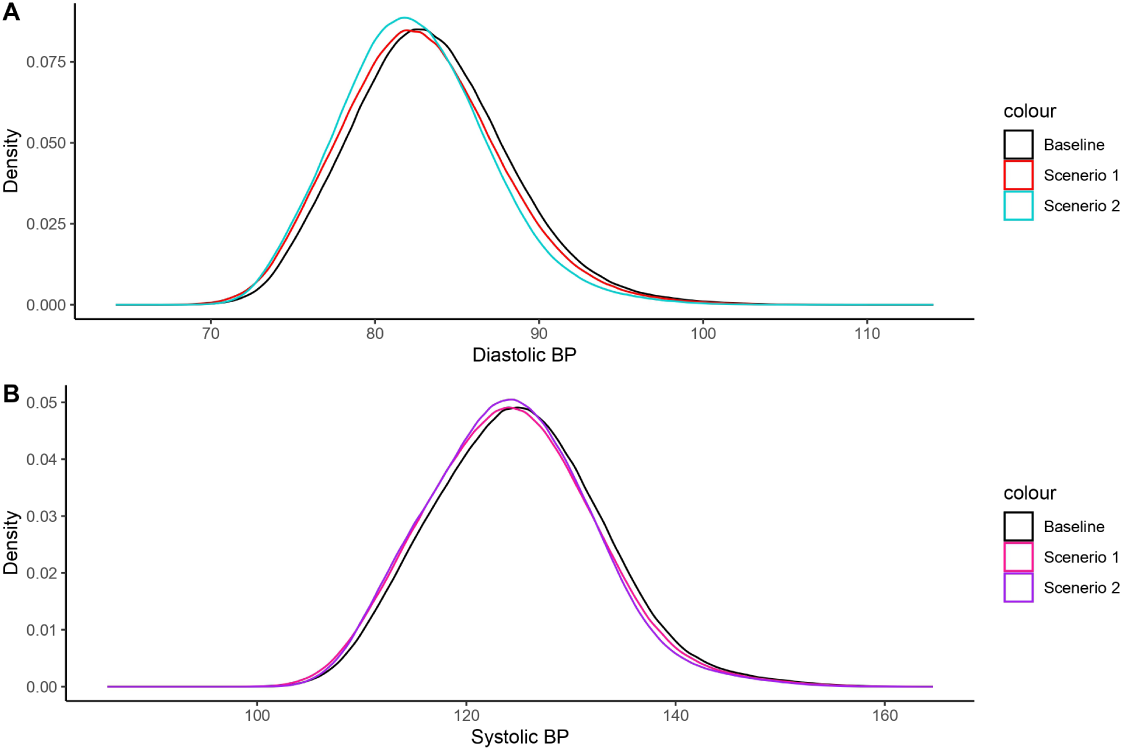
Distribution of systolic and diastolic blood pressure under baseline, scenario 1, and scenario 2 simulations.

Table 8 displays the hypertension prevalence within the simulated population across three scenarios: baseline, Scenario 1, and Scenario 2. In Scenario 1, the hypertension prevalence decreased to 16.15% from 19.17% in the baseline simulation. This suggests that a one-unit decrease in mean BMI at age 20 has the potential to lead to a 3% decline in hypertension prevalence within the population, assuming the relationship equation between age, BMI, and blood pressure remains the same. In Scenario 2, the hypertension prevalence was found to be 15.33%, which is 3.8% lower than the baseline simulation model. This implies that if the standard deviation of BMI at age 20 were one unit lower, the hypertension prevalence would have decreased by 3.8% in the population, again assuming the relationship equation between age, BMI, and blood pressure remains constant.

**Table 8:** Prevalence of hypertension under baseline, scenario 1 and scenario 2 simulations.

|  | Baseline | Scenerio1 | Scenerio2 |
| --- | --- | --- | --- |
| Urban-Male | 31.11 | 27.21 | 26.21 |
| Urban-Female | 22.13 | 18.66 | 17.73 |
| Rural-Male | 15.71 | 12.57 | 11.89 |
| Rural-Female | 10.19 | 8.28 | 7.74 |
| Total Population | 19.17 | 16.15 | 15.33 |

Notably, the largest decrease in the prevalence of hypertension was observed among urban males, with a decline of 3.9% in scenario 1 and a decline of 4.9% in scenario 2. Conversely, the lowest decline in prevalence was observed among rural females, with a decrease of 1.9% in scenario 1 and 2.5% in scenario 2.

Table 9 presents the changes in hypertension prevalence among individuals aged 50 or older across the three scenarios: baseline, scenario 1, and scenario 2. This table holds particular significance due to the sharp increase in hypertension prevalence observed after the age of 50, in conjunction with the aging population in India. In the baseline simulation, the prevalence of hypertension for individuals aged 50 or older was recorded at 28.7%. However, in scenario 1, this prevalence decreased to 25.7%, indicating a reduction in hypertension incidence. Moreover, in scenario 2, the prevalence of hypertension further decreased to 24.6%. Notably, at age 70, the prevalence of hypertension was found to be 38.8% in scenario 1, which is 5% lower than the prevalence observed in the baseline simulation. This suggests that if there are two cohorts of individuals aged 20 years, with the second cohort having a mean BMI one unit lower than the first cohort and the same standard deviation, the prevalence of hypertension at age 70 is expected to be 5% lower in the second cohort compared to the first cohort.

**Table 9:** Prevalence of hypertension among individuals aged 50 or above under Scenario 1 and Scenario 2.

|  | Baseline | Scenerio1 | Scenerio2 |
| --- | --- | --- | --- |
| Age $\geq 50$ | 28.71 | 25.69 | 24.61 |
| Age 70 | 43.81 | 38.77 | 36.31 |

Additionally, in Scenario 2, the prevalence of hypertension at age 70 was found to be 36.9%, which is 6.9% lower than the hypertension prevalence observed at age 70 in the baseline model. This implies that if there are two cohorts of individuals aged 20 years, with the second cohort having one unit less standard deviation in BMI distribution and the same mean, the prevalence of hypertension at age 70 would be approximately 7% lower in the second cohort compared to the first cohort.

## 4 Discussion

This study employs dynamic microsimulation modelling to investigate the potential impact of reducing obesity and modifying the distribution of BMI on hypertension prevalence. The findings indicate that, following the current BMI distribution, mortality rates, and the relationship between age, BMI, and blood pressure, a cohort of 100,000 individuals aged 20 will exhibit an obesity prevalence of 11.4% and hypertension prevalence of 43.8% at age 70. Viewing the cohort experiences from a cross-sectional perspective indicates a BMI prevalence of 10.3% and hypertension prevalence of 19.2% in the simulated population. The study highlights that a one-unit decrease in the mean value of BMI at baseline (age 20) would lead to an approximately 3% reduction in hypertension prevalence in the simulated population. Conversely, a one-unit reduction in the standard deviation of BMI at baseline would result in a 3.8% decrease in hypertension prevalence in the simulated population. However, it is crucial to interpret these findings with consideration of the modelling assumptions and limitations. While acknowledging the crude nature of the model used in this study, it offers insights into the significance of targeting obesity for hypertension reduction.

When comparing the two scenarios - a one-unit decrease in mean BMI, and a one-unit decrease in the standard deviation of BMI, the higher reduction in prevalence of hypertension was observed under scenario-2, involving the reduction in the spread of BMI distribution at baseline. These findings indirectly point towards the necessity of addressing the spread of BMI distribution in the population. The current mean BMI values are not inherently problematic; rather, the concern lies with the extremes of the BMI distribution. This suggests the importance of targeting individuals with both high and low BMI values at young ages by providing accurate information about nutrition and healthy eating habits. By doing so, not only will the spread of the BMI distribution decrease, but the mean of the distribution will also be reduced. As emphasized in prior research, it is worth noting that not only has the average BMI been on the rise in recent decades, but the variance of BMI has also experienced a significant increase [14]. This observation underscores the importance of considering both the mean and variance of BMI when identifying the most effective intervention strategies [14].

The findings of this study indirectly underscore the importance of addressing childhood obesity. The global prevalence of childhood obesity has markedly risen over the past three decades, particularly in developed nations [15]. While current levels of childhood obesity in India remain relatively low, proactive measures are essential to prevent a similar upward trajectory. Research suggests that children classified as overweight or obese are prone to carrying these conditions into adulthood, increasing their susceptibility to non-communicable diseases such as diabetes and cardiovascular ailments at a younger age [16]. The profound impact of childhood obesity extends to physical health, social and emotional well-being, and self-esteem, with lasting effects persisting into later life [16]. Therefore, it is important for the policymakers and the stakeholders to understand the gravity of this issue and formulate viable strategies to address and mitigate its consequences.

The findings of this study must be interpreted with due consideration for the limitations, as well as the validity and reliability of the modelling employed. It is crucial to note that the microsimulation model utilized in this study does not simulate the entire population; rather, it provides a microsimulation of 100,000 individuals aged 20 years, projecting their experiences based on current patterns of mortality and BMI over the next 50 years. Essentially, the model predicts the outcome at age 70 if this specific cohort follows the given parameters of BMI, blood pressure, and mortality. However, it is important to highlight that the interpretation of the results are made at a population level, assuming that the experiences of individuals at each age are indicative of cross-sectional outcomes. Another limitation lies in the adjustment of the association equation between BMI and hypertension, which was solely done for age within all four sex-residence groups. The model’s scope for refinement could have been enhanced by incorporating numerous other factors into the equation. However, the model’s inherent structure restricts the consideration of additional factors.

Several factors can influence the pattern and distribution of BMI across different age groups. These may encompass variables such as sex, residence (urban or rural), socioeconomic status, education level, region, and genetic factors, po-tentially contributing to heterogeneity in BMI distribution. However, due to data constraints, our model could only incorporate sex and residence. While the input data provided separate probabilities of death for all four sex-residence groups based on life tables from SRS, we were unable to adjust these probabilities of death based on BMI level and hypertension status. For instance, consider two rural females aged 50 years: one with hypertension and a BMI of 27.0, and the other without hypertension with a BMI of 23.0. Ideally, the probability of death should differ significantly between the two individuals, with the first woman having a higher probability of death compared to the second one. However, in our model, both individuals would have the same probability of death because we lacked data on the probability of death categorized by hypertension status and BMI level. These limitations present opportunities for improvement in future studies addressing this topic. Future studies can incorporate additional life events into the simulation framework. These events could encompass key life milestones such as marriage, childbirth, and employment, providing a more comprehensive representation of individuals’ life trajectories. Additionally, including risk of other morbidities with increase in age would contribute to a more nuanced understanding of health dynamics within the simulated population.

While it is crucial to acknowledge the limitations of this study, it is equally important to recognize its significant contribution to the existing body of literature. It contributes to the relatively limited body of literature on the application of microsimulation modelling in health research within the context of India. In developed countries, the reliance on microsimulation modelling to shape fiscal and economic policies is well-established. European Union countries, for instance, utilize EUROMOD, a tax-benefit microsimulation model, to analyse the effects of taxes and benefits on household incomes. Similarly, the United States employs models like DYNASIM3, MINT, and CBOLT, Canada uses DYNACAN, Norway employs MOSART, Sweden utilizes SESIM, Australia relies on APPSIM, and the UK utilizes SAGE and PENSIM microsimulation models to analyse diverse outcomes within their respective populations. These models play an active role in policymaking by providing insights into questions that are challenging to address using alternative modelling techniques. For instance, these models have been instrumental in exploring various scenarios related to future old age pension schemes and their potential impact on both micro and macroeconomic aspects [17].

While numerous countries are using microsimulation models to shape their policies and interventions, there is almost no attention is being paid on this type of models in India. There is a need for increased research focus and funding dedicated to microsimulation modelling in India. Although the development of a robust microsimulation model at the national or subnational level is a prolonged and resource-intensive process demanding substantial funding and a skilled research workforce, the outcomes it yields are undeniably valuable and worth investment.

## Data Availability

All the data used in the study is publicly available

## Appendix

**Table A1:** 95% bound of change in BMI from previous age.

|  | Urban-Male |  | Urban-Female |  | Rural-Male |  | Rural-Female |  |
| --- | --- | --- | --- | --- | --- | --- | --- | --- |
|  | LB | UB | LB | UB | LB | UB | LB | UB |
| **Age** |  |  |  |  |  |  |  |  |
| 21 | 0.082 | 0.571 | 0.288 | 0.341 | 0.104 | 0.229 | 0.281 | 0.330 |
| 22 | 0.036 | 0.487 | 0.333 | 0.375 | 0.140 | 0.264 | 0.270 | 0.323 |
| 23 | 0.045 | 0.426 | 0.347 | 0.398 | 0.191 | 0.317 | 0.265 | 0.325 |
| 24 | 0.088 | 0.389 | 0.349 | 0.398 | 0.171 | 0.294 | 0.250 | 0.303 |
| 25 | 0.121 | 0.356 | 0.347 | 0.393 | 0.162 | 0.268 | 0.249 | 0.291 |
| 26 | 0.161 | 0.353 | 0.340 | 0.378 | 0.157 | 0.248 | 0.240 | 0.273 |
| 27 | 0.183 | 0.352 | 0.326 | 0.358 | 0.158 | 0.241 | 0.233 | 0.262 |
| 28 | 0.195 | 0.354 | 0.311 | 0.337 | 0.142 | 0.223 | 0.217 | 0.244 |
| 29 | 0.198 | 0.345 | 0.288 | 0.316 | 0.116 | 0.198 | 0.201 | 0.230 |
| 30 | 0.161 | 0.296 | 0.265 | 0.293 | 0.093 | 0.180 | 0.183 | 0.214 |
| 31 | 0.152 | 0.274 | 0.242 | 0.278 | 0.073 | 0.164 | 0.166 | 0.198 |
| 32 | 0.152 | 0.266 | 0.218 | 0.260 | 0.065 | 0.159 | 0.150 | 0.188 |
| 33 | 0.107 | 0.221 | 0.192 | 0.239 | 0.048 | 0.144 | 0.131 | 0.173 |
| 34 | 0.073 | 0.182 | 0.176 | 0.231 | 0.025 | 0.124 | 0.113 | 0.159 |
| 35 | 0.042 | 0.137 | 0.158 | 0.226 | -0.004 | 0.097 | 0.094 | 0.144 |
| 36 | 0.014 | 0.115 | 0.135 | 0.221 | -0.015 | 0.086 | 0.085 | 0.137 |
| 37 | 0.006 | 0.112 | 0.114 | 0.215 | -0.036 | 0.064 | 0.079 | 0.140 |
| 38 | 0.000 | 0.111 | 0.093 | 0.208 | -0.057 | 0.041 | 0.066 | 0.134 |
| 39 | 0.008 | 0.125 | 0.071 | 0.199 | -0.058 | 0.037 | 0.053 | 0.125 |
| 40 | 0.020 | 0.138 | 0.046 | 0.183 | -0.055 | 0.035 | 0.043 | 0.123 |
| 41 | 0.040 | 0.157 | 0.046 | 0.191 | -0.059 | 0.028 | 0.035 | 0.130 |
| 42 | 0.052 | 0.163 | 0.041 | 0.200 | -0.065 | 0.020 | 0.036 | 0.136 |
| 43 | 0.052 | 0.170 | 0.031 | 0.204 | -0.068 | 0.009 | 0.025 | 0.131 |
| 44 | 0.057 | 0.187 | 0.031 | 0.221 | -0.068 | 0.029 | 0.008 | 0.117 |
| 45 | 0.043 | 0.176 | 0.021 | 0.229 | -0.052 | 0.054 | -0.024 | 0.099 |
| 46 | 0.020 | 0.147 | 0.019 | 0.238 | -0.028 | 0.078 | -0.052 | 0.081 |
| 47 | 0.029 | 0.149 | 0.014 | 0.241 | -0.016 | 0.100 | -0.081 | 0.063 |
| 48 | 0.032 | 0.155 | -0.012 | 0.231 | -0.031 | 0.095 | -0.109 | 0.033 |
| 49 | 0.037 | 0.177 | -0.019 | 0.243 | -0.049 | 0.088 | -0.123 | 0.021 |
| 50 | 0.042 | 0.194 | -0.049 | 0.229 | -0.059 | 0.090 | -0.134 | 0.017 |
| 51 | 0.041 | 0.191 | -0.073 | 0.223 | -0.061 | 0.097 | -0.131 | 0.025 |
| 52 | 0.060 | 0.210 | -0.098 | 0.215 | -0.085 | 0.088 | -0.119 | 0.038 |
| 53 | 0.025 | 0.236 | -0.130 | 0.200 | -0.104 | 0.078 | -0.114 | 0.043 |
| 54 | -0.014 | 0.227 | -0.130 | 0.207 | -0.123 | 0.065 | -0.106 | 0.043 |
| 55 | -0.062 | 0.203 | -0.172 | 0.172 | -0.150 | 0.043 | -0.083 | 0.061 |
| 56 | -0.100 | 0.181 | -0.203 | 0.146 | -0.168 | 0.042 | -0.068 | 0.070 |
| 57 | -0.086 | 0.231 | -0.200 | 0.156 | -0.184 | 0.051 | -0.070 | 0.068 |
| 58 | -0.086 | 0.252 | -0.201 | 0.163 | -0.186 | 0.072 | -0.082 | 0.056 |
| 59 | -0.109 | 0.249 | -0.190 | 0.185 | -0.171 | 0.108 | -0.095 | 0.038 |
| 60 | -0.092 | 0.283 | -0.197 | 0.199 | -0.177 | 0.120 | -0.123 | 0.005 |
| 61 | -0.117 | 0.277 | -0.201 | 0.226 | -0.156 | 0.158 | -0.127 | 0.000 |
| 62 | -0.140 | 0.277 | -0.225 | 0.244 | -0.163 | 0.174 | -0.137 | -0.001 |
| 63 | -0.173 | 0.290 | -0.255 | 0.268 | -0.167 | 0.203 | -0.155 | -0.004 |
| 64 | -0.239 | 0.296 | -0.301 | 0.283 | -0.164 | 0.257 | -0.160 | 0.015 |
| 65 | -0.333 | 0.308 | -0.352 | 0.282 | -0.195 | 0.301 | -0.172 | 0.031 |
| 66 | -0.452 | 0.308 | -0.415 | 0.252 | -0.262 | 0.317 | -0.180 | 0.051 |
| 67 | -0.452 | 0.390 | -0.496 | 0.188 | -0.358 | 0.275 | -0.188 | 0.056 |
| 68 | -0.424 | 0.411 | -0.669 | 0.103 | -0.449 | 0.210 | -0.163 | 0.090 |
| 69 | -0.921 | 0.552 | -1.224 | 0.187 | -0.971 | 0.211 | -0.282 | 0.208 |
| 70 | -0.921 | 0.552 | -1.224 | 0.187 | -0.971 | 0.211 | -0.282 | 0.208 |

LB: lower bound; UB: upper bound

**Table A2:**
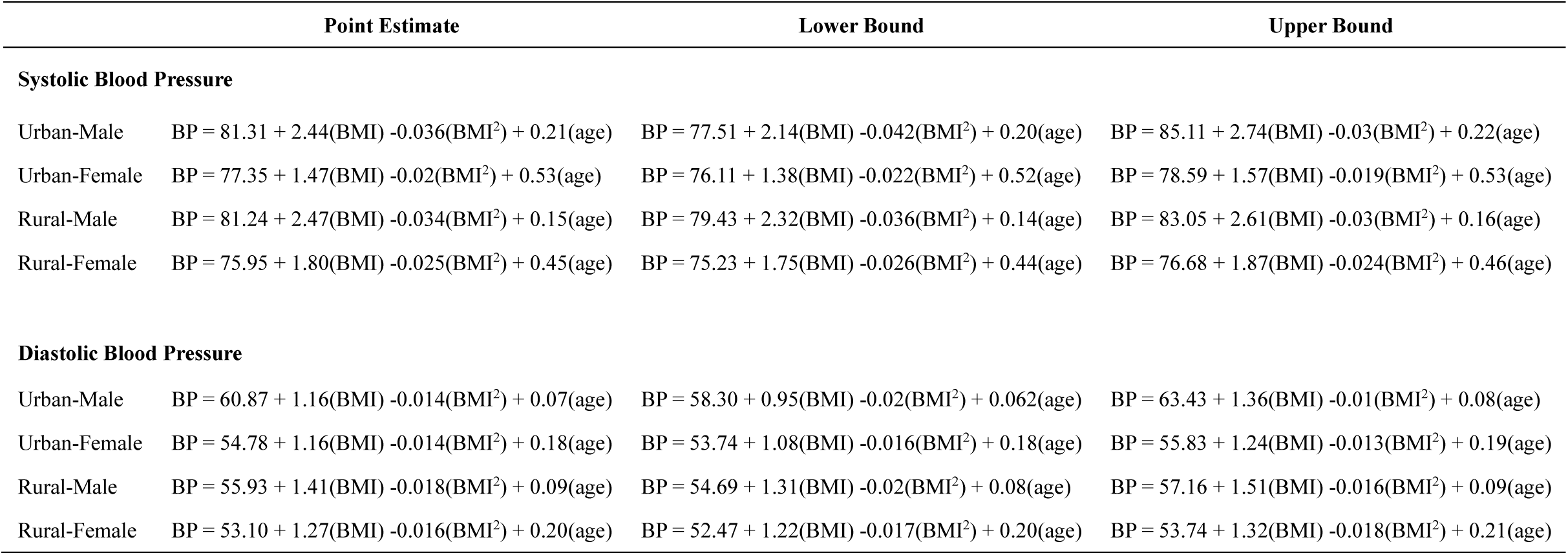
Equations of association between BMI, Age, and Blood Pressure.

